# The Association of Lymphocyte count and levels of CRP, D-Dimer, and LDH with severe coronavirus disease 2019 (COVID-19): A Meta-Analysis

**DOI:** 10.1101/2020.04.20.20072801

**Authors:** Almigdad H. M. Ali, Sagad Omer Obeid Mohamed, Ibrahim H. E. Elkhidir, Mohamed Elata Hassan Elbathani, Abazr A. H. Ibrahim, Almutasim B. E. Elhassan, Mohammed Suliman Tawer Salman, Mazin A.M. Elhassan, Mahmoud Elnil, Abdelhamid Ibrahim Hassan Abuzied

## Abstract

The rapid progression of Coronavirus disease 2019 (COVID-19) and its increasing burden on health systems necessitate the identification of parameters of severe infection to help in monitoring, prognoses and development of treatment algorithms. This review aims to investigate the association of lymphocyte count, CRP, LDH, and D-Dimer with the severe form of COVID-19. This review was conducted according to the Preferred Reporting Items for Systematic Reviews and Meta-Analyses (PRISMA) guidelines. The databases of MEDLINE/PubMed, WHO-Virtual Health Library (VHL), and ScienceDirect were used for the systematic search. Random effects model was used to estimate the pooled standardized mean differences (SMD) with the corresponding 95% confidence interval (CI), using OpenMeta Analyst software. A total of 11 studies, with 2437 COVID-19 patients, which fulfilled the eligibility criteria were included in the meta-analysis. The analysis revealed that lymphocyte count was significantly lower in patients with the severe form of COVID-19 (SMD = - 1.025, P value <.001). Also, the analysis of SMD showed that patients with severe COVID-19 have a significantly higher serum levels of CRP (SMD = 3.363, P value <.001), D-Dimer (SMD = 1.073, P value <.001), and LDH (SMD = 3.345, P value <.001). Low lymphocyte count and high levels of CRP, LDH, and D-Dimer are associated with severe COVID-19. These laboratory markers could be used as clinical indicators of worsening illness and poor prognosis of COVID-19.

## Introduction

In December 2019, Wuhan - the capital of Central China’s Hubei province-had witnessed the emergence of a new virus of the *Coronaviridae* family that caused several cases of severe respiratory disease [1-2]. The virus was named by the world health organization (WHO) as Severe Acute Respiratory Syndrome novel Coronavirus 2 (SARS-CoV-2) and the disease caused by this virus as Coronavirus Disease 2019 (COVID19) [1-2]. The disease kept spreading dramatically to reach other provinces in China and other countries around the world, forcing the WHO to declare it as a global pandemic on the 11^th^ of March 2020 [3]. Despite the efforts done to contain the disease, it has spread to over 200 countries around the world, causing a significant burden at the level of health facilities, as well as economic burdens on international systems [4].

The SARS-CoV-2 is transmitted by infective respiratory droplets from infected patients, fomites, and surfaces reaching conjunctiva or respiratory mucosa and the incubation period usually ranges from 2 to 14 days after exposure, during which the patient may be infectious. [4-5]. Animal to human transmission is still being investigated, knowing that similar coronaviruses were found in bats, rodents and birds [5-6].

Most people infected with COVID-19 experienced a mild to moderate respiratory illness, with cough and shortness of breath being the commonest symptoms [6-7]. Approximately 14% of the patients developed a severe disease with complications like acute respiratory distress syndrome and respiratory failure, acute liver injury, acute kidney injury, cardiovascular complications, septic shock, and multi-organ failure [6-7]. Older people and those with underlying co-morbidities like diabetes, chronic respiratory diseases, cardiovascular diseases, and immune-compromised patients had a higher risk of developing severe disease [8].

Patients with severe disease or complications require special care such as admission to intensive care units (ICU) with mechanical ventilation [9]. However, there is a wide gap between the total population and the number of intensive care beds available [9]. Further adding to the burden, ICU services cost the hospitals around 39% of the total drug costs, 25% of the equipment, and 13% of the lab investigations [10]. Researchers are still working to find a definitive treatment of COVID-19 and there are ongoing trials on some antiviral, anti-inflammatories, and anti-malarial drugs such as hydroxyl-chloroquine sulfate and chloroquine phosphate products to be used for certain hospitalized patients with COVD-19 [5, 11].

Diagnosis is dependent on the evaluation of the risk of contact with an infected patient, clinical signs and symptoms, and polymerase chain reaction testing of viral RNA on respiratory samples [1, 4-5]. Other investigations have been used to evaluate the patient’s condition and to predict severe outcomes of the disease such as complete blood count, D-Dimer, Procalcitonin, C-reactive protein (CRP), and lactate dehydrogenase (LDH) [4-5]. Two recent meta-analyses showed that increased Procalcitonin values and thrombocytopenia are associated with a higher risk of severe COVID-19 [12-13]. Other studies suggested that lymphocytes count, D-Dimer, CRP, and LDH have a role in the prognosis of COVID-19. However, and to the best of our knowledge, there is no meta-analysis on these biomarkers. Therefore, the objective of this review is to assess the association between severe COVID-19 and these laboratory markers. Taking into consideration the disease burden and the limited resources and capacities of health facilities, it is very crucial to find parameters that support the clinical condition’s risk assessment to aid the anticipation of severe complications.

## Materials and Methods

### Search strategy and inclusion criteria

In this meta-analysis, we followed the Preferred Reporting Items for Systematic Reviews and Meta-Analyses (PRISMA) statement [14]. The systematic literature search was performed using the electronic databases of Medline/PubMed, WHO-Virtual health library (VHL), and ScienceDirect, without date or language restriction. The search terms used were (Novel coronavirus), (2019 nCoV), (COVID-19), (SARS-CoV-2), (Lymphopenia), (Lymphocytopenia) (leukopenia), (leukocytopenia), (D-Dimer), (C-reactive protein), (CRP), (lactate dehydrogenase), and (LDH) to ensure no possible relevant articles were missed. Also, we reviewed the articles referenced by those identified articles in this search.

All observational studies reporting sufficient information on lymphocyte count, CRP, LDH, and D-Dimer levels in both severe and non-severe COVID-19 patients, with a clear definition of severe illness, were included in the analysis for calculation of the standardized mean difference (SMD) estimates. The exclusion criteria were: case reports, editorials, letters, abstracts, and studies without sufficient data of interest. If two or more studies had the same patient population, the study with more complete data was included to avoid duplication.

The titles and abstracts of all articles retrieved from this search were screened for potential inclusion in this review. Then, potentially relevant studies were reviewed (full text) for inclusion according to the defined eligibility criteria, and the data were extracted by independent reviewers using a data extraction form. The quality of the studies was assessed using the Newcastle – Ottawa scale. Any disparity among the reviewers at any step was resolved by discussion and consensus.

### Data analysis

The statistical analyses were carried out by OpenMeta Analyst software version 10.10 for analysis [15]. The pooled SMD was calculated from the random-effects model due to the notable heterogeneity in this meta-analysis. Statistical heterogeneity was assessed with the I^2^ statistics and publication bias was determined through Egger’s test and visual examination of the funnel plot [16].

## Results

### Characteristics of the studies

The schematic flow of the studies identification and selection process is presented in (Figure 1) and the summary of the data from the included studies is shown in (Table 1). Our search retrieved records for 285 published articles. Of which, full texts of 34 potentially relevant studies were retrieved for full-text screening and 23 studies were subsequently omitted because of duplication of the study populations and insufficient data to estimate the outcomes of interest. Lastly, a total of 11 studies with 2437 patients, published from December 2019 to March 2020 were included for the analysis. All of them were from China, Mainland [17-26], whilst one study was based in Singapore [27].

**Figure 1:**
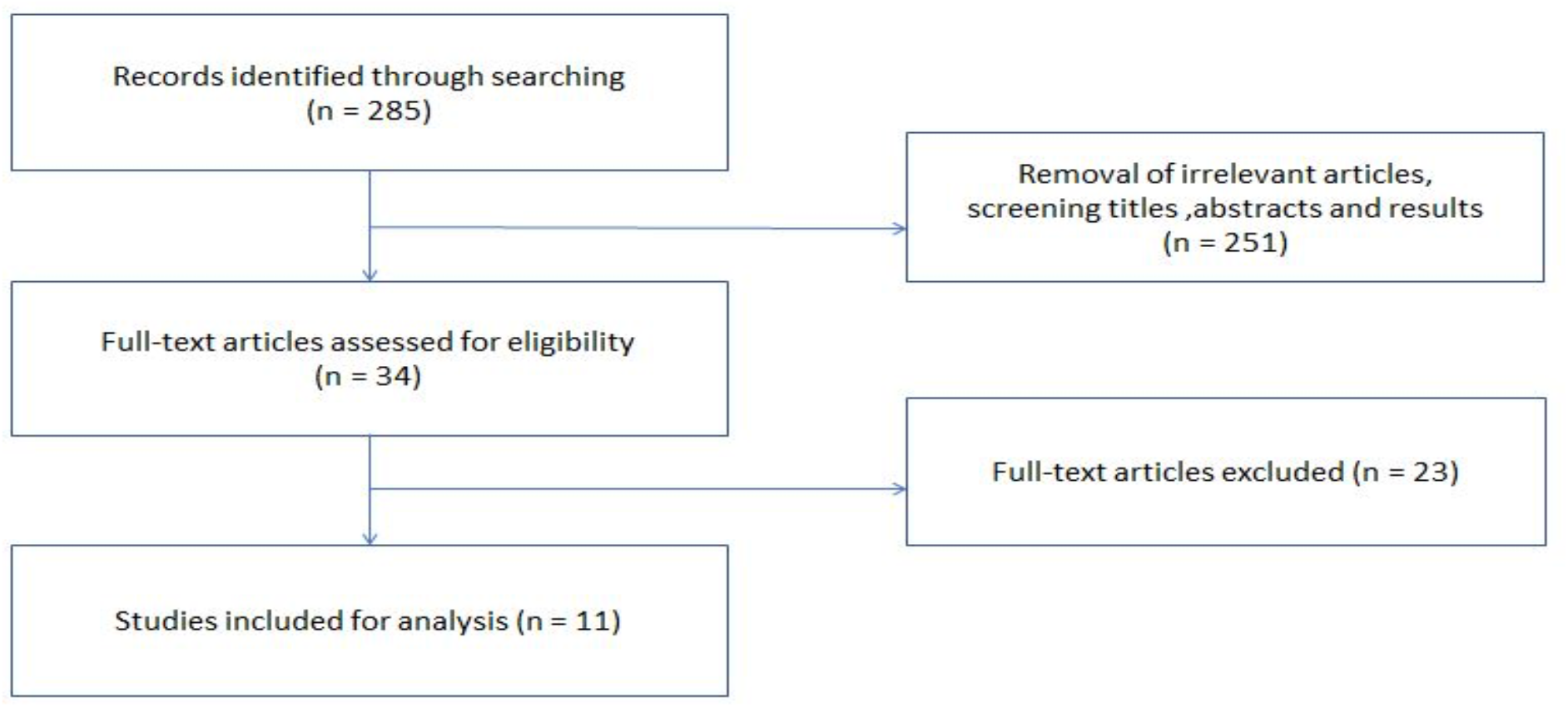
The flow diagram for the process of study selection and systematic review of literature.

Severe COVID-19 was determined based on the composite of respiratory distress with hypoxia and/or hypoxemia in three studies [17-19], the American Thoracic Society guidelines for community-acquired pneumonia in one study [20], Youden’s index in one study [21], guidelines of expert Chinese group in one study [22], non-survival in one study [23], disease progression in one study [24], Oxygen saturation < 90% in one study [25], the requirement of ICU admission in one study [26], and the requirement of supplemental Oxygen in one study [27].

### Analysis of SMD of the lymphocyte count between the two groups of patients

The pooled effect size showed that the lymphocyte count was significantly lower in patients with severe COVID-19 than patients with non-severe COVID-19 and the SMD = - 1.025 (95% CI, -1.336 – -0.714: P<.001) (Figure 2). No evidence of publication bias was detected based on visual examination of the funnel plot and from the results of Egger’s test (p = 0.59) (figure 6 A).

**Figure 2:**
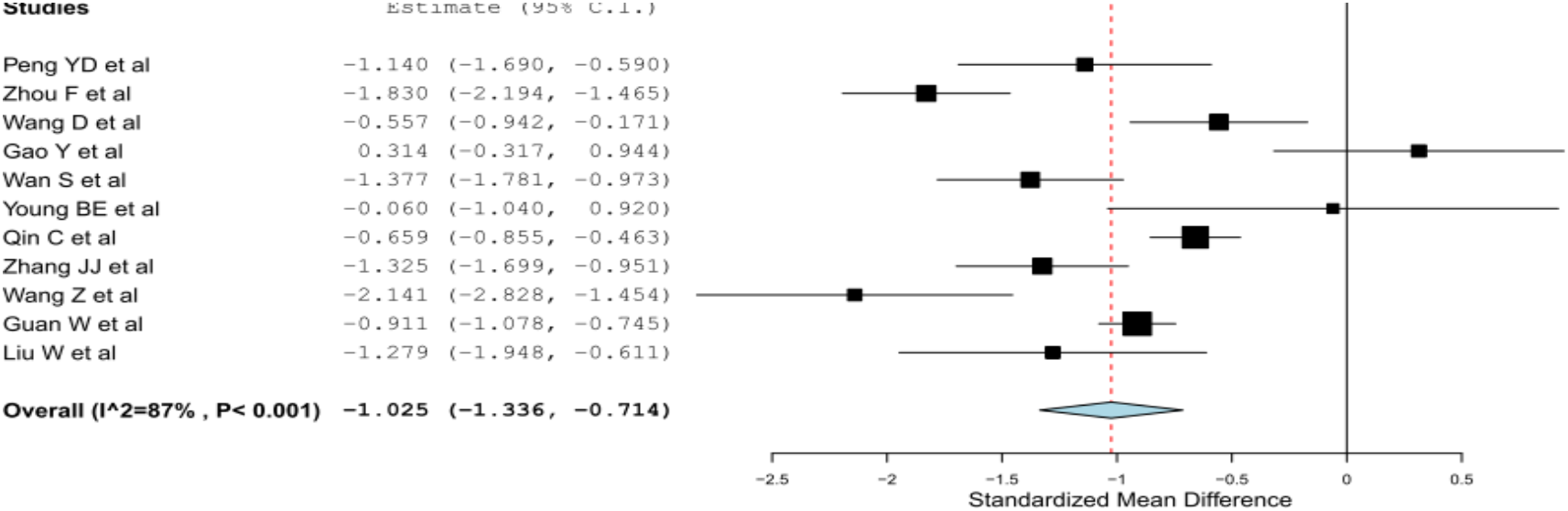
Pooled SMD of lymphocyte count estimates between the two groups of patients (severe and non-severe COVID-19)

### Analysis of SMD of the CRP between the two groups of patients

The pooled effect size showed that CRP level was significantly higher in patients with severe COVID-19 than patients with non-severe COVID-19, with an SMD = 3.363 (95% CI, 2.203 – 4.522: P<.001) (Figure 3). We detected a slight publication bias was detected based on visual examination of the funnel plot and from the results of Egger’s test (p = 0.03) (figure 6 B). The Duvall and Tweedie trim and fill method indicated that potential missing studies were three.

**Figure 3:**
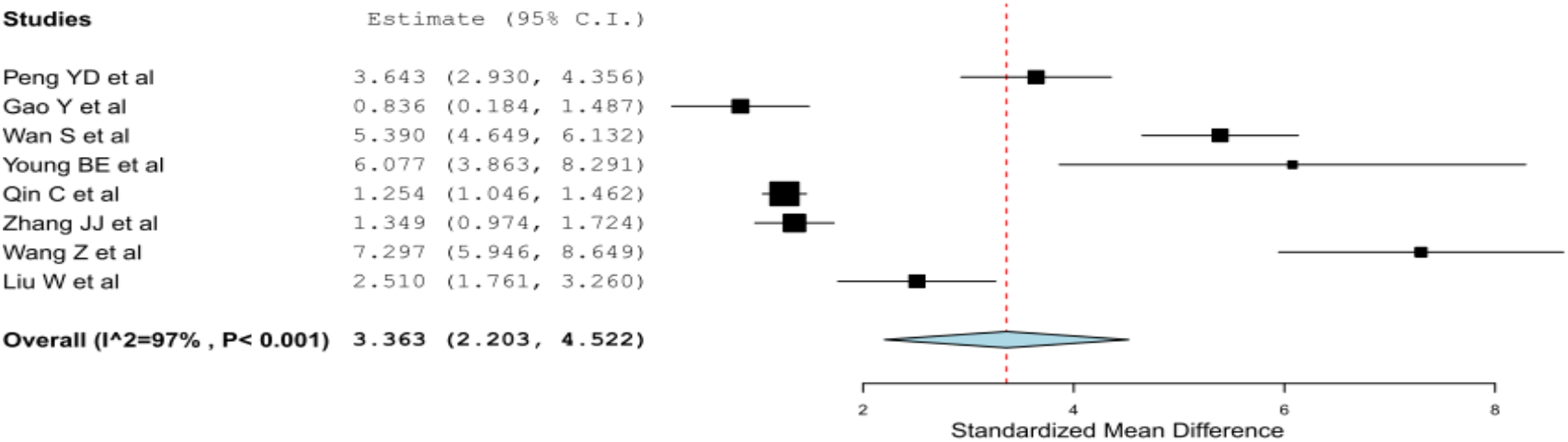
Pooled SMD of CRP estimates between the two groups of patients (severe and non-severe COVID-19)

### Analysis of SMD of the D-Dimer between the two groups of patients

The pooled effect size showed that D-Dimer level was significantly higher in patients with severe COVID-19 than patients with non-severe COVID-19, with an SMD = 1.073 (95% CI, 0.728 – 1.418: P<.001) (Figure 4). No evidence of publication bias was detected based on visual examination of the funnel plot and from the results of Egger’s test (p = 0.97) (figure 6 C).

**Figure 4:**
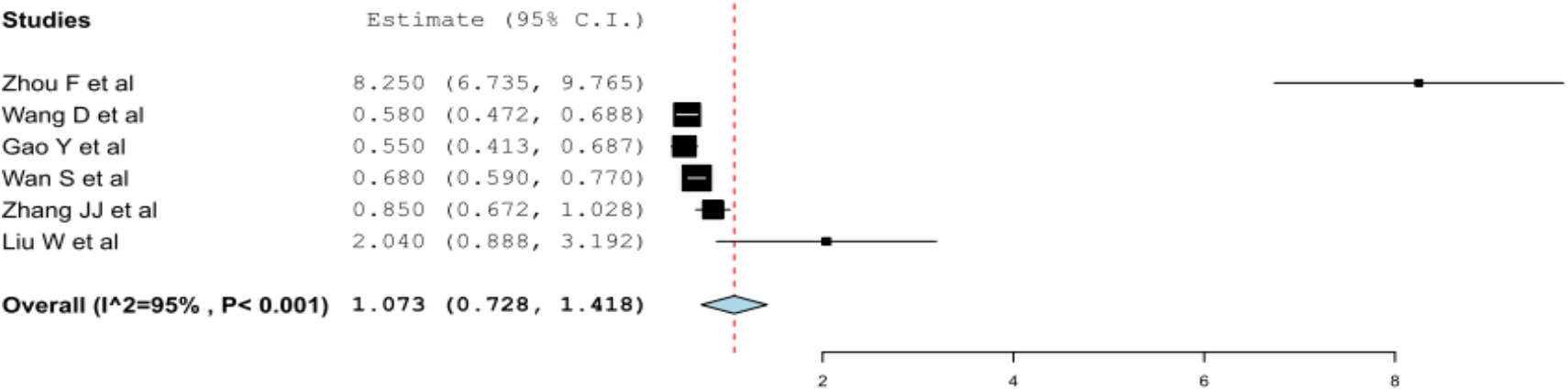
Pooled SMD of D-Dimer estimates between the two groups of patients (severe and non-severe COVID-19)

### Analysis of SMD of the LDH between the two groups of patients

The pooled effect size showed that LDH level was significantly higher in patients with severe COVID-19 than patients with non-severe COVID-19, with an SMD = 3.345 (95% CI, 1.925 – 4.765: P<.001) (Figure 5). No evidence of publication bias was detected based on visual examination of the funnel plot and from the results Egger’s test (p = 0.83) (figure 6 D). The leave-one-out sensitivity analyses indicated that our results were robust and were not driven by any single study.

**Figure 5:**
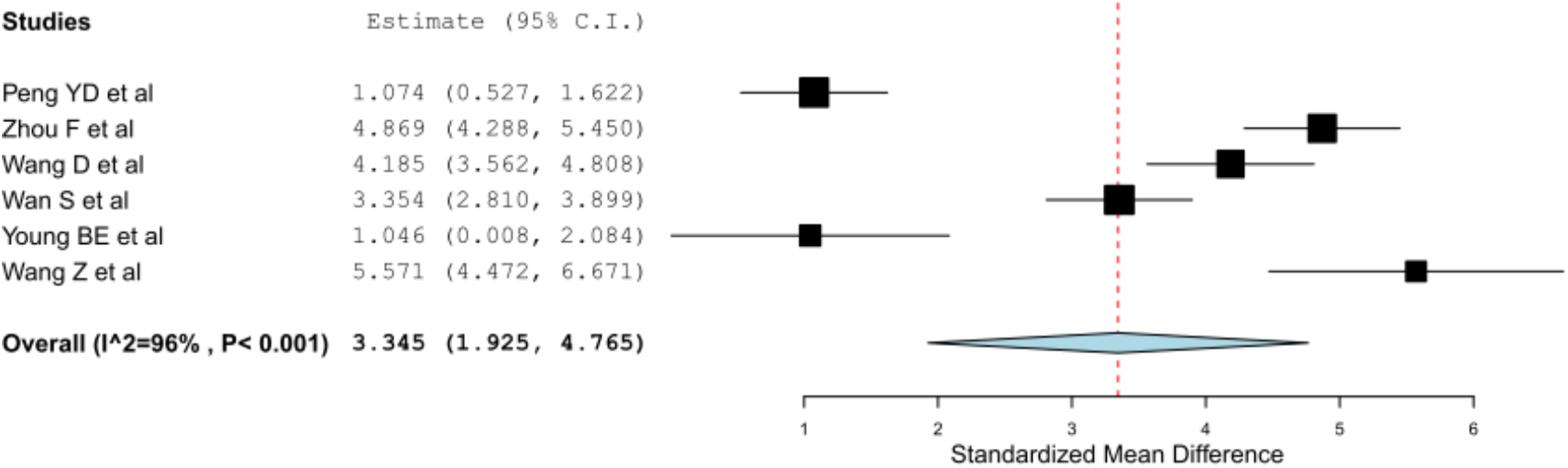
Pooled SMD of LDH estimates between the two groups of patients (severe and non-severe COVID-19)

**Figure 6(A-D):**
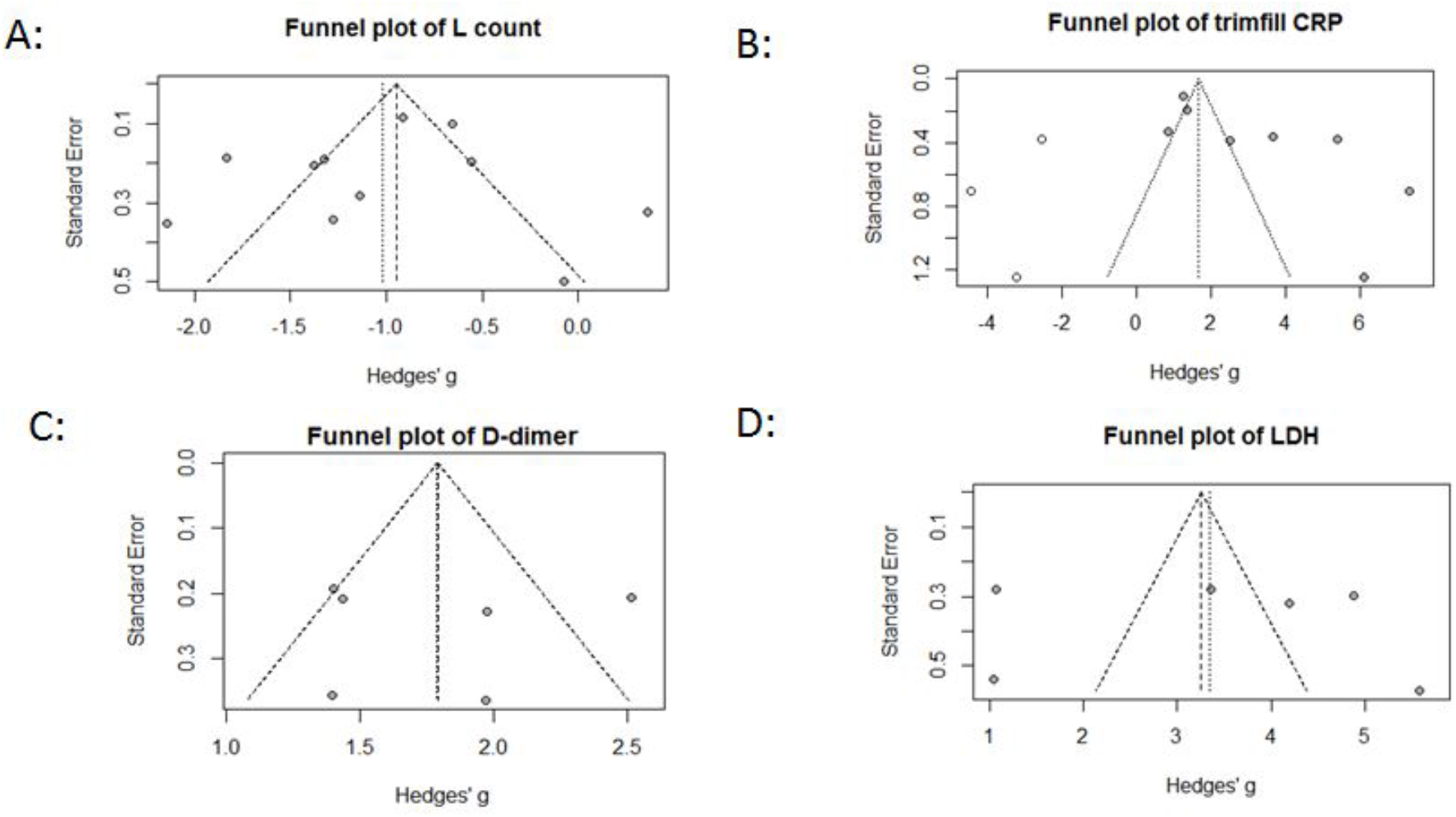
Funnel plots for assessment of the publication bias.

## Discussion

In addition to the clinical criteria, it is of great benefit to have laboratory markers for severe infection which will help in monitoring and prognosis and will be used to develop workup and treatment algorithms. This meta-analysis investigated the association between severe COVID-19 and several biomarkers.

The analysis showed that patients with poor outcomes of COVID-19 are likely to have lymphopenia as shown in (figure 2). Based on several studies, it has been suggested that the immune system is impaired during the disease and COVID-19 might damage T lymphocytes [6, 8, 28]. These studies showed a particular decrease in the CD4+ subset of lymphocytes and higher naive CD4+ cells than memory cells in severe cases [6, 8, 28]. The higher naive : memory cells ratio indicates that the immune system was impaired more severely [6, 8]. The resulted cytokine storm in the body generates a series of immune responses that cause changes in peripheral white blood cells [28].

The levels of CRP and other inflammatory markers, such as IL-6, IL-8, IL-2R, and IL-10, were noticed to be higher in severe cases than non-sever cases [22]. Increased levels of cytokines, chemokines, and Neutrophils to Lymphocyte Ratio (NLR) in severe cases suggest a possible hyper-inflammatory response role in the pathogenesis of COVID-19 [22]. The D-Dimer level elevation in patients with severe disease may raise the suspicion of underlying abnormal blood coagulation function [22].

The effect of D-Dimer widens to include not only the correlation with the severity of the disease but also with mortality percentage, as shown by Zhou F. et al study in Wuhan, China, who found a D-Dimer level of more than 1 microgram/ml is associated with fatal outcome [23]. Gao Y. et al. showed that the clinical benefit of D-dimer level prediction of the severity of the disease will increase if it is combined along with IL6 level [21]. Also, the level of LDH, which is an inflammatory marker, shows a similar correlation with mortality percentages as that of D-dimer [21, 23].

The findings of this study need to be considered in the context of some limitations. All studies that we found were in Asia, whereas nowadays the majority of cases are in the United States and Europe. Due to the limitedness of the available data and variable definition of disease severity among the studies, there is a need for further studies to explore and clarify the prognostic role, as well as the precise mechanisms underlying the changes in these biomarkers in patients with severe COVID-19.

## Data Availability

Available upon request

## Ethical approval and consent to participate

not applicable

## Consent for publication

not applicable.

## Availability of data and material

The dataset generated during this study are available from the corresponding author on reasonable request.

## Authors contribution

(AA) conceptualized the research idea; (AA, SM, and IE) designed the study; (AA, SM, ME, MS, AI and AE,) undertook articles searching, articles assessment, and data extraction; (SM, and IE) undertook data analysis; All authors interpreted the results and drafted the manuscript. All authors revised and approved the final manuscript.

## Acknowledgment

Non to acknowledge

## Supplementary Materials

Table 1: Summery of the included studies in the review.

## Abbreviations

COVID-19: Coronavirus Disease 19
SARS-Cov-2: Severe Acute Respiratory Syndrome novel Coronavirus 2
CRP: C-Reactive Protein
LDH: Lactate Dehydrogenase

